# Membrane Computing Simulation of Sexually Transmitted Bacterial Infections in Hotspots of Individuals with Various Risk Behaviors

**DOI:** 10.1101/2023.07.03.23292134

**Authors:** Marcelino Campos, Juan Carlos Galán, Mario Rodríguez-Dominguez, José M. Sempere, Carlos Llorens, Fernando Baquero

## Abstract

The epidemiology of sexually transmitted infections (STIs) is complex due to the coexistence of various pathogens, the variety of transmission modes derived from sexual orientations and behaviors at different ages and genders, and sexual contact hotspots resulting in network transmission. There is also a growing proportion of recreational drug users engaged in high-risk sexual activities, as well as pharmacological self-protection routines fostering non-condom practices. The frequency of asymptomatic patients makes it difficult to develop a comprehensive approach to STI epidemiology. Modeling approaches are required to deal with such complexity. Membrane computing is a natural computing methodology for virtual reproduction of epidemics under the influence of deterministic and stochastic events with an unprecedented level of granularity. The application of the LOIMOS program to STI epidemiology illustrates the possibility of using it to shape appropriate interventions. Under the conditions of our basic landscape, including sexual hotspots of individuals with various risk behaviors, an increase in condom use reduces STIs in a larger proportion of heterosexuals than in same-gender sexual contacts and is much more efficient for reducing *N. gonorrhoeae* than *Chlamydia* and lymphogranuloma venereum infections. Amelioration from diagnostic STI screening could be instrumental in reducing *N. gonorrhoeae* infections, particularly in men having sex with men (MSM), and *C. trachomatis* infections in the heterosexual population; however, screening was less effective in decreasing lymphogranuloma venereum infections in MSM. The influence of STI epidemiology of sexual contacts between different age groups (<35 and ≥35 years) and in bisexual populations were also submitted for simulation.

**Importance:** The epidemiology of sexually transmitted infections (STIs) is complex and significantly influences sexual and reproductive health worldwide. Gender, age, sexual orientation, sexual behavior (including recreational drug use and physical and pharmacological protection practices), the structure of sexual contact networks, and the limited application or efficiency of diagnostic screening procedures creates variable landscapes in different countries. Modeling techniques are required to deal with such complexity. We propose the use of a simulation technology based on membrane computing, mimicking *in silico* STI epidemics under various local conditions with an unprecedented level of detail. This approach allows us to evaluate the relative weight of the various epidemic drivers in various populations at risk and the possible outcomes of interventions in particular epidemiological landscapes.

## INTRODUCTION

In the first decade of the 21st century, the incidence of sexually transmitted infections (STIs) increased worldwide, with an even more marked increase in recent years. Thus, all international organizations consider STIs a current public health problem that requires a rapid and effective multidisciplinary approach. In 2020, the World Health Organization (WHO) estimated 374 million new STIs, with bacterial infections caused by *Chlamydia trachomatis* and *Neisseria gonorrhoeae* the most frequently detected (https://www.who.int/news-room/fact-sheets/detail/sexually-transmitted-infections). These infections have considerable impact on the sexual and reproductive health of infected people, both symptomatic and asymptomatic (1vg). The 75th Assembly of the WHO, highly aware of the worldwide increase in STIs, redesigned the Global Health Sector Strategy from 2016-2021, including in the same program HIV, hepatitis, and STIs for the 2022-2030 period (2wh). This integrated view could be an excellent opportunity to include STIs in national surveillance programs, which have examined other diseases with a high burden and population impact. Two aspects of concern are particularly relevant: the worldwide underdiagnosis of STIs as a consequence of poor screening and low risk perception from individuals and groups prone to exposure and infection.

Recently, the Australian Government Department of Health, through the Kirby Institute, published their 2021 national STI incidence. In accordance with data from international organizations, they found a significant increase in gonorrhea and a slight increase in *Chlamydia*. However, they estimate that only 20%-28% of all gonorrhea and *Chlamydia* infections have been diagnosed (3kj). Moreover, there are large differences (around 100-fold) in the incidence of *Chlamydia* infections among various European countries, probably due to differences in the amount of chlamydia testing (4eu). Unfortunately, this situation is compounded by the fact that many infected people have no signs or symptoms of infection (5fe). In our experience, in Spain, 80% of *C. trachomatis* infections and 20% of LGV infections are asymptomatic (data not shown). Infected asymptomatic individuals and those who do not perceive their risk of infection are key to the maintenance of STI infections in the population.

Another factor influencing STI epidemiology is derived from highly interconnected sexual networks, which facilitate the rapid spread of new infections among people with high-risk sexual behaviors (e.g., multiple sexual partners, condomless sex, sex with unknown people, alcohol use, recreational drug use), revealing the excessive vulnerability to new threats, as we learned with the recent international mpox outbreak in 2022 (6hj), which is associated with sexual transmission (7ck). High connectivity is facilitated by online dating apps for recreational sex, which expand sexual networks (8hm). The increase in reported cases that are closely associated with highly interconnected networks could also be due in part to the progressive implementation of pre-exposition prophylaxis (PrEP) to prevent new HIV infections in people with high exposure and infection risk. To improve the fight against the HIV pandemic, the PrEP strategy has also generated less condom use (9aa). Since PrEP implementation, a reduction in new HIV diagnoses has coincided with an increase in STIs (10aa).

The overwhelming complexity of factors determining STI epidemiology, involving various microbial organisms, modes and frequencies of transmission derived from sexual orientations and behaviors at different ages, the use of chemsex (drugs facilitating or enhancing sexual activity), PrEP, the variable symptomatology along the infective process (eventually treated), and the difficulties in detecting asymptomatic cases, makes it difficult to provide an integrated view of this significant health problem, which can only be approached by using mathematical or computational modeling. Although simulation modeling to understand STI spread remains scarcely implemented, recent international public health emergencies decreed by the WHO in 2022 (concerning mpox) have stimulated this approach. Its recent use in the international mpox outbreaks has contributed to understanding and predicting STI evolution (11si). In this study, we simulate STI transmission patterns by membrane computing, which derives from the larger field of cellular computing (12pj, 13pa). Membrane computing offers a unique opportunity to mimic epidemiological scenarios including individuals with different susceptibility to STI exposure (due to their age, gender, sexual orientation), which are represented as individual entities defined as “membranes.” These membranes are exposed by the action of particular “objects”; for example, particular types of protective tools (condoms), drugs or, in our case, microbes involved in STIs. These individuals can interact and transmit STIs in a particular meeting location and according to specific sexual behaviors, with variable probabilities of contact and transmission, in accord with rules that can be defined in the simulation model. Such interactions occur inside larger “environmental” membranes, describing the places where sexual meetings take place. In short, membrane computing creates scenarios in which events occur according to local conditions, probabilities, and the intensity of interactions between computational entities, resulting in predictable outcomes. In practical terms, cellular membrane computing mimics reality, creating “virtual epidemics” in the model, according to variant (à la carte) parameters established by the researcher, which can be based on the observed reality. Thus, various STI-oriented interventions (single or combined) can be applied to specific scenarios as a guide to select the most efficient. Most importantly, the model can help to identify possible unknown variables or parametric data (which are frequently difficult or impossible to obtain) influencing epidemic outcomes and can include the new variable and/or refine the parameters in such a way that the model adjusts to the local observed data. We have previously explored membrane computing applications in clinical microbiology in the fields of antibiotic resistance epidemiology and in SARS CoV-2 epidemic events and their vaccination control (14mc, 15mc, 16mc, 17gg). Our simulator (which is applicable to any infectious disease) is named LOIMOS, from the ancient Greek *loimos* (*λιμ*ό*ς*), meaning plague, pestilence, or any deadly infectious disorder. A user-friendly interface is being developed for LOIMOS, and it will be freely available. Interested readers should contact our first author, Dr. Marcelino Campos, for more information.

## METHODS: THE BASIC EPIDEMIOLOGICAL PARAMETERS OF THE MODELED SCENARIO

### Outlook of the model: general demographics, distribution, and mobility

In this study, we modeled STIs spreading within a target population of 16,000 sexually active people (8000 males and 8000 females) of various ages seeking sexual encounter hotspots. Each gender has 2000 heterosexuals (males seeking sex with females/females seeking sex with males [MSF/FSM]; for simplicity in the text, heterosexuals are termed MSF) in each age group (<35 and ≥35 years) and 2000 homosexuals (males seeking sex with males [MSM] or females seeking sex with females [FSF] in equivalent numbers) in each age group. The present model aims to reveal the hypothetical dynamics of STI transmission among sexual hotspot users. Although it is difficult to generalize gender and sexual orientation proportions, and numerical differences could bias the results for a given group of users, the model allows the researcher to modify these proportions. In this model, the population lives in 20 home residential areas (HRAs) that differ in the density of inhabitants. The target sex-seeking population moves between home and various sexual encounter hotspots or “sexual exchange hotspots” (SEHs) where they can have sexual relations, eventually resulting in potential infection and STI transmission. These SEHs are distributed across 3 SEH networks (20 zones per network), corresponding to the preferential types of sexual partners: MSM, FSF, and MSF, creating distinct SEH-*n* locations corresponding to these 3 networks (e.g., SEH-01-MSM, SEH-01-FSF, SEH-01-MSF; SEH-02-MSM, SEH-02-FSF, SEH-02 MSF, and so on). In the basic model, half of the SEHs in each network are predominantly visited by those <35 years of age, and the other half by older individuals. Each individual can visit from 0-3 SEHs per day. In each network, we distinguished 14 mobility patterns (MPs), expressing the probability for an individual to visit a particular SEH within their network. For instance, individuals moving according to MP-06 have a 50% probability of going to SEH-06 and a 50% probability of going to SEH-07 in his/her network. If the individual following MP-06 is an MSF, he will visit SEH-06 and SEH-07 of the MSF network with these probabilities. If he is an MSM, he will visit SEH-06-MSM and SEH-07-MSM of the MSM network with these probabilities. In one of the variants of the basic scenario presented in this study, an individual from one age group visits a SEH corresponding to younger or older individuals with a probability of 10%. In the following sections we present the epidemiological parameters used in the basic model; however, we insist that most of these values can be modified by the user to mimic specific local situations. A depiction of the theoretical model is presented in Figure 1.

**Figure 1.**
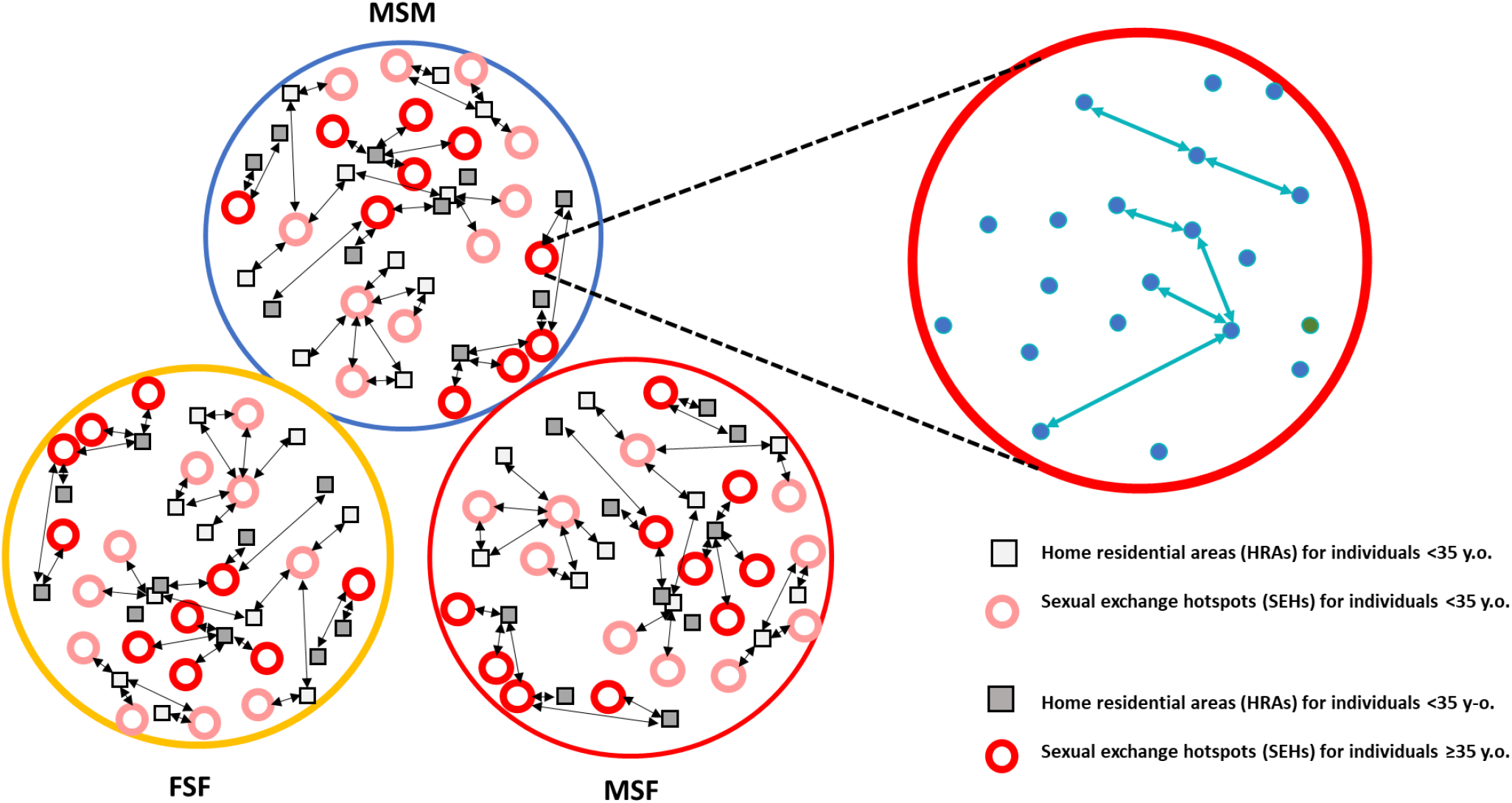
Schematic representation of the theoretical simulation’s basic model. The 3 circles on the left correspond with the 3 basic sexual contact networks determined by sexual preference (MSF, males having sex with females; MSM, males having sex with males; and FSF, females having sex with females). Black arrows indicate the movements from residential areas (HRAs) to sexual exchange hotspots (SEHs). On the right, a detail of a particular SEH (corresponding to MSM ≥35) with the individuals represented in blue dots, and their sexual interactions in green arrows.

### Epidemiological parameters for *C. trachomatis* infection and lymphogranuloma venereum used in the basic model

**In individuals of <35 years of age**, in *symptomatic patients* (having symptoms during the infection period), there is an incubation period of approximately 10 days; during this period, the patient does not transmit the infection to other partners. Next, the patient has 4 pre-symptomatic days (days 10-14 after contagion), followed by mild symptoms lasting for 12 days, during which the infected person can transmit the microorganism; then comes a period of moderate-severe symptomatology lasting for 15 days, during which sexual activity is substantially decreased or discontinued and contagion does not take place. When the patient is under therapy (30 days for *C. trachomatis* or lymphogranuloma venereum [LGV]) and in the post-therapy period (10 days) we do not consider any possibility of contagion to other individuals. Globally, we estimate that in patients aged <35 years there is a 40% probability of having an asymptomatic disease (19pc). In these *asymptomatic patients*, the non-contagious phase of incubation is also 10 days, followed by 75 days during which, because of the absence of symptoms, contagion is possible. The natural disappearance of contagion occurs within the following 10 days.

**In individuals ≥35 years of age,** there is a 40% probability of asymptomatic infection. In the *symptomatic patients*, after a 10-day non-contagious incubation period, the individual enters in a pre-symptomatic period for 5 days (days 10-15), followed by a phase with mild symptoms lasting for 15 days, during which the patient can potentially transmit the *C. trachomatis* infection. Then, there is a period of moderate-severe symptomatology lasting for 20 days, during which sexual activity is substantially decreased or suspended, and contagion to other individuals does not take place. When the patient is diagnosed and receives therapy (30 days, see above), and in the post-therapy period (10 days) there is no contagion. *In asymptomatic patients*, the early non-contagious phase (incubation period) is also 10 days, followed by 100 days during which patients are unaware of their infection and contagion is possible. Within the successive 10 days, when the infection naturally disappears, the individual becomes non-contagious.

Parameter ranges consider the European STI epidemiology (18gm), which is based on diagnosed cases; the number of asymptomatic cases can only be estimated. These data indicate higher peak *C. trachomatis* infections among women <35 (approximately 340/100,000) than in males of this age group (approximately 160/100,000). In Europe, LGV cases, caused by invasive *C. trachomatis* serotypes, are almost exclusively detected in MSM (approximately 35/100,000, slightly more in those ≥35 than in younger individuals). For the purposes of this study, our basic model is focused on the *C. trachomatis* STI in the group of MSF <35, whereas LGV is considered associated with the MSM population ≥35.

### Epidemiological parameters for *N. gonorrhoeae* infection used in the basic model

In the case of *N. gonorrhoeae* infections, we distinguish epidemiological parameters between individuals <35 and those ≥35, male and female, and considering their sexual orientation. As in the former case of *C. trachomatis* infections, a critical point in the epidemiology is the proportion of symptomatic versus asymptomatic cases.

**In the male population aged <35** infected by *N. gonorrhoeae*, we considered in our basic model that asymptomatic cases account for 20% of MSF, 40% of bisexual MSF/MSM, and 55% of MSM. In these asymptomatic patients, and after the 3-day incubation period, the patient is contagious for the next 38 days. Contagion does not occur from 10 days following the spontaneous cure of the infection.

*Symptomatic cases* have a 3-day incubation period, 4 pre-symptomatic non-contagious days, then 4 days during which symptoms are mild and contagion could take place. During the next period, with typical symptomatology lasting for 4 days, the infected person does not transmit the infection because of the suspension of sexual activity. Standard therapy consists of a single intramuscular administration of cephalosporin. During the period in which antibiotic exposure reduces the symptomatology of the infection (11 days) sexual activity might occur; however, during this time and in the next post-antibiotic period (10 days), the patient is not considered contagious.

**In the male population ≥35 years of age**, the proportion of asymptomatic cases for the various sexual orientations is the same as in the population <35 (see above). ***Symptomatic cases***, following a 3-day incubation period, enter into a contagious period encompassing 5 pre-symptomatic days and subsequently a 5-day mildly symptomatic phase. When typical symptoms appear, and during the next 5 days, contagion drops because of a strong reduction of sexual activity. Under the antibiotic effect influencing the evolution of the infection (next 2 weeks), and for the following 10 days, the patient is not considered contagious. In ***asymptomatic cases***, after 3 days of incubation, contagion from the patient to other individuals can occur during the next 50 days. Within 10 days after the natural resolution of the infection, the individual becomes non-contagious.

**In the female population <35 years of age,** we consider a high (80%) frequency of asymptomatic infection. *Symptomatic patients* have a 10-day incubation period, followed by 7 days of early-phase, pre-symptomatic infection during which transmission can occur. During the following 7 days of mild symptomatology the patient is also contagious; when signs and symptoms are evident, and for 11 days, sexual activity is strongly reduced or suspended and transmission does not take place. During the period of antibiotic effect (14 days), and in the following 10 days, the patient is not considered contagious. *In asymptomatic patients*, after 10 days of incubation time and during the following 45 days we consider that contagion to other individuals can take place; contagion ceases 10 days after spontaneous local clearance of the infection.

**In the female population ≥35 years of age,** as in the <35 group, there is an 80% probability of asymptomatic infection. In *symptomatic patients*, after a 10-day incubation period, the infected individual enters an 8-day pre-symptomatic phase and then an 8-day mildly symptomatic period; during these 16 days the patient is contagious. When typical symptoms appear, sex is discontinued for 14 days; thus, the probability of transmission is near 0%. During antibiotic therapy (14 days) and 10 days after therapy there is no risk of contagion. In the *asymptomatic patients*, after the 10 days of incubation there is a prolonged period (60 days) during which the patient could transmit the infection, ending with the local clearance of *N. gonorrhoeae;* this non-contagious period begins within 10 days following infection clearance.

Epidemiological parameter ranges consider the European STI epidemiology, based on diagnosed *N. gonorrhoeae* infection cases; the number of asymptomatic cases can only be estimated. Considering sexual orientation, we estimated a prevalence of infection among the male population <35 years of 170/100,000; in males ≥35 of 70/100,000; in females <35 of 70/100,000; and in females ≥35 of 20/100,000. In our basic model, we focus on heterosexual individuals <35 and MSM <35.

### STI co-occurrence in the model scenario

In our current model, for simplicity, we consider that a patient who has acquired an STI (*C. trachomatis* or *N. gonorrhoeae*) cannot be infected again by the same microorganism during the simulation period. However, this possibility exists in the clinical setting (different *C. trachomatis* clones) and could be considered in further versions of our membrane computing models. In the present version, we consider that for a given STI in a symptomatic patient, the sexual interactions are suppressed and therefore they cannot acquire any new STIs.

### Populations with protected sex, chemsex, and/or PrEP in the STI model scenario

Protected sex refers to sexual activity during which a condom is used to protect against STIs. In our model, there is protected sex if either of the 2 sexual partners use condoms by their own initiative. We have explored 3 levels of protected sex: high-level usage (95% of sexual encounters), medium-level usage (50%), and low-level usage (5%). The probability of STI contagion without protection was estimated for both genders at 22% for *C. trachomatis* and LGV. In *N. gonorrhoeae*, these probabilities are 55% for males and 38.5% for females. For both infections, the probability of contagion while using protection was always 5%.

Chemsex is the consumption of recreational drugs to facilitate or enhance sexual activity, such as methamphetamine, paramethoxyamphetamine, ketamine, mephedrone, gamma-hydroxybutyrate, cocaine, ecstasy, amyl-nitrite, and other compounds. PrEP consists of drugs taken to prevent HIV infection (most frequently tenofovir disoproxil-fumarate, lamivudine, and emtricitabine), which is associated with decreased condom use. In our model scenario, the proportion of only chemsex users is 23.6%; of daily PrEP 3%; of punctual (only in particular occasions) PrEP 1%; of PrEP plus chemsex 4.3% (1.7% if punctual PrEP); and the rest, non-users of either PrEP or chemsex 66.4%. In summary, the total proportion of chemsex users was 29.6%, and of PrEP 10%.

Visiting a SEH is frequently associated with the decision to use chemsex and/or PrEP. We considered in the simulation model that such usage would increase the possibility of acquiring an STI by 25% in chemsex consumers, with or without PrEP, and approximately 20% in punctual PrEP users, without chemsex. Among individuals not using PrEP or chemsex, high-medium-low protection frequency was estimated to occur in 40%, 30%, and 30%, respectively. In the case of individuals with early infection belonging to the bisexual, continuous PrEP user, and chemsex user groups, the high-medium-low sexual protection levels were 30%, 30%, and 40%, respectively. Considering the entire population, the high-medium-low protection user rate was 36.60%, 29.93%, and 33.48%, respectively (mean data from various parallel simulations). Integrating all modeled values for each sexual encounter, there was an expected possibility of using protection in 76.4% of cases, and having sex without protection in 23.6%.

The reduction in the infected population by protection depends on the STI and gender of the infected patient. For *C. trachomatis* infection, the contagion probability with protection is 5% of sexual encounters; without protection, 22% in both males and females. For *N. gonorrhoeae* infection, in males the contagion probability with and without protection is 5% and 55%, respectively; in females, the corresponding figures are 5% and 38.5%, respectively. Note that sexual encounters outside sexual hotspots have an estimated risk for STI infection of 0.001% between partners of the same age group and 0.0000001% between individuals belonging to different age groups. Individuals not visiting sexual hotspots were not considered in this study.

### Individual host mobilities for visiting sexual encounter hotspots in the STI model scenario

In our simulation we differentiated 4 out-of-home mobility patterns (groups) for hosts seeking sex, according to the relative proportion of individuals and the daily possibility of such mobility during the week, considering a choice of 3 possible mobilities (with sexual contacts) per day. Group 1 (30% of the individuals) had a 40% possibility of mobility from Monday to Thursday, 80% on Friday, and 80% over the weekend; group 2 (50% of individuals) had a 20%, 60%, and 60% probability, respectively; group 3 (10% of individuals) a 0%, 0%, and 10% probability, respectively; and group 4 (10% of individuals) a 60%, 20%, and 20% probability, respectively. Chemsex users had an additional 20% possibility of visiting a SEH. In other words, we considered in the model that each host had 3 daily attempts to leave the home to seek sex. For instance, an individual belonging to group 3 had a 10% probability during the weekend of leaving home to seek sex on the first day and a 10% probability of doing so on the second or third occasion on the same day; thus, if this host succeeded at each attempt they had a probability of 30% to accomplish *at least* one move seeking out-of-home sex over the week-end.

Our simulated population is located in 20 various HRAs, differing in the density of individuals visiting sexual hotspots. Therefore, the target sex-seeking population can leave home to visit 60 different sexual encounter hotspots or SEHs where they can have sexual contact, possibly resulting in STI transmission. From each of the HRAs the individual can, according to their sexual orientation, attend a particular SEH (any of the 20) constituting a network for heterosexuals, 20 for male and 20 for female same gender sexual behavior). The probability of visiting a particular SEH is the product of 2 consecutive decisions: first, to attend a SEH; second, which to select, depending on sexual orientation (see Table 1). We consider in the model a total of 20 SEHs in each of the networks defined by the preferential type of sexual partner. Of these, the first half are mostly visited by hosts <35 and the second half by those ≥35. We can include cross-mobility among zones, defined by 2 main patterns: 1) 10% of the time, those <35 visit older individuals’ zones; and 2) 10% of the time, those ≥35 visit younger individuals’ zones. Such variation was not included in the basic model, only in a particular variant of it (see the specific section below). Each of these patterns was studied in 3 separate networks, corresponding to heterosexuals and same-gender sex males and females (Figure 1). In Table S1 (Supplementary Material) we show 14 mobility patterns (MPs), representing the probabilities that a given individual from a given residency area will access a particular SEH in 20 different residencies in town (HRAs). We considered in the model that individuals sharing the same HRA would have similar mobility patterns (MPs) to neighbor SEHs. Conversely, contagion between neighbors of the same residency area (out of SEHs) occurs with much less probability. For example, in MP5, the individual in this residency area has a 25% probability of visiting SEH 4, 50% probability of visiting SEH 5, and 25% probability of visiting SEH 6. The simulated number and shared profiles of hosts from each residency (mobility pattern, residential area, sexual orientation, gender, and age group) are detailed in Table S2. For instance, for *C. trachomatis* infection, we started in HRA-20 (MSF) with 10 infected individuals <35, 5 males and 5 females; for LGV, HRA-1 (MSM) had 10 infected individuals ≥35; for *N. gonorrhoeae*, HRA-11 (MSM) had 10 infected individuals <35; and in HRA-14 (MSF), there were 10 MSFs, with both genders equally represented.

## RESULTS

### Evolution of STI numbers after introduction of infected people in sexual networks: basic scenario

Figure 2 shows the evolution of contagion numbers over time for the 3 types of sexual networks as determined by sexual preference (MSM, FSF, and MSF), which are also subdivided according to age. In the graphs, each day is represented by 3 steps in the abscissa, so that 90 steps correspond roughly to 1 month, and 1095 to a year. In total, 5000 steps are represented, approximately 4.5 years. The sexual behavior group with the highest number of cases is, as expected, the MSM <35 group (1300 cases for LGV, 1200 for *N. gonorrhoeae*, and 900 for *C. trachomatis*).

**Figure 2.**
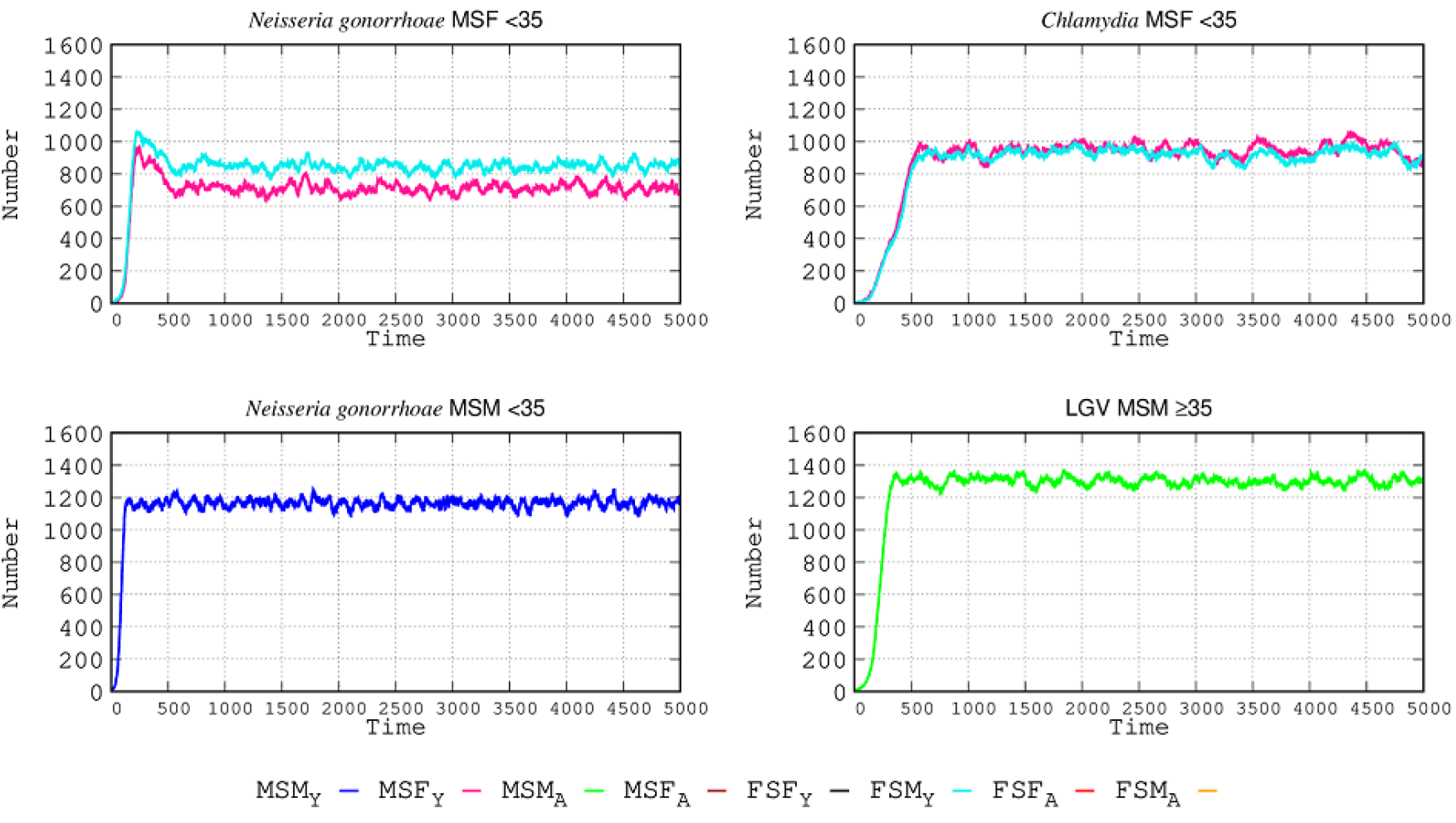
STI epidemiology in the simulated populations. Top panels: in the MSF group, males are represented in by the red line and females by the light blue line. Lower panels: in the MSM group the dark blue line corresponds to men <35 and green line to men ≥35. Only the groups having infected cases are represented.

Given that the sexual networks were separated by age in our simulation, there was no cross contagion. The increase in contagion occurs more rapidly in *N. gonorrhoeae* than in *C. trachomatis.* In the MSF population <35, the frequency of infected females <35 (light blue line) surpasses that of the males <35 (red line), which can be attributed to more frequent sub-symptomatic infection, and also the increased length of infection in comparison with males, both symptomatic and asymptomatic. The initial peak in *N. gonorrhoeae* is reduced once *C. trachomatis* infections reach a maximum number, which is higher than in *N. gonorrhoeae*; patients with gonorrhea who acquire *C. trachomatis* reduce their sexual activity and become less frequently infected. This is because, even if females have a higher proportion of asymptomatic cases, it is compensated for by males who more frequently have sex without protection. The frequency of gonorrheal disease among MSM <35 is much higher (58%) than in the case of MSF <35. In the case of LGV, the number of infected individuals (all MSM ≥35) was even higher than the number of those with gonorrheal disease in MSM <35, and the slope of the epidemic curve was steeper than for *C. trachomatis infection* in the MSF <35 population. Once a maximum number of cases was reached, the number of infections leveled off over time. Given that the MSF <35 group was exposed to 2 STIs (*N. gonorrhoeae* and *Chlamydia*), the number of infected patients for each of the STIs is lower that that in the MSM <35 and MSM ≥35 groups.

### The effect of increased STI protection tools

In this new scenario, we increased the proportion of individuals using STI protection tools (such as condoms). High-level usage increased from 36.60% to 56.40%, medium-level usage was reduced from 29.93% to 19.92%, and low-level usage decreased from 33.48% to 23.68%. Considering all values, in this new scenario there is an 87.55% probability per sexual encounter of using protection (12.45% non-using). The effect of protection changes with the various STIs and sexual behaviors is shown in Figure 3, where the effect of the increase in protection tools at step 5000 is represented. In MSF interactions this increase reduces the *N. gonorrhoeae*-infected patients by 35% in females and 43% in males <35. Reduction in the MSM population <35 was lower, at 21%. For *C. trachomatis* infection, in sexual relations among MSFs <35, the reduction is much lower than in the case of *N. gonorrhoeae*, at only 20%. The number of lymphogranuloma cases in males ≥35 was only reduced by 10% after the increase in protective tools. For both types of *Chlamydia* infections, the reduction in the number of cases after an intervention with higher protection takes longer to occur than in the case of *N. gonorrhoeae*, due to the higher transmission rate of *N. gonorrhoeae*: the higher the transmission rate, the greater the effectiveness of protection.

**Figure 3:**
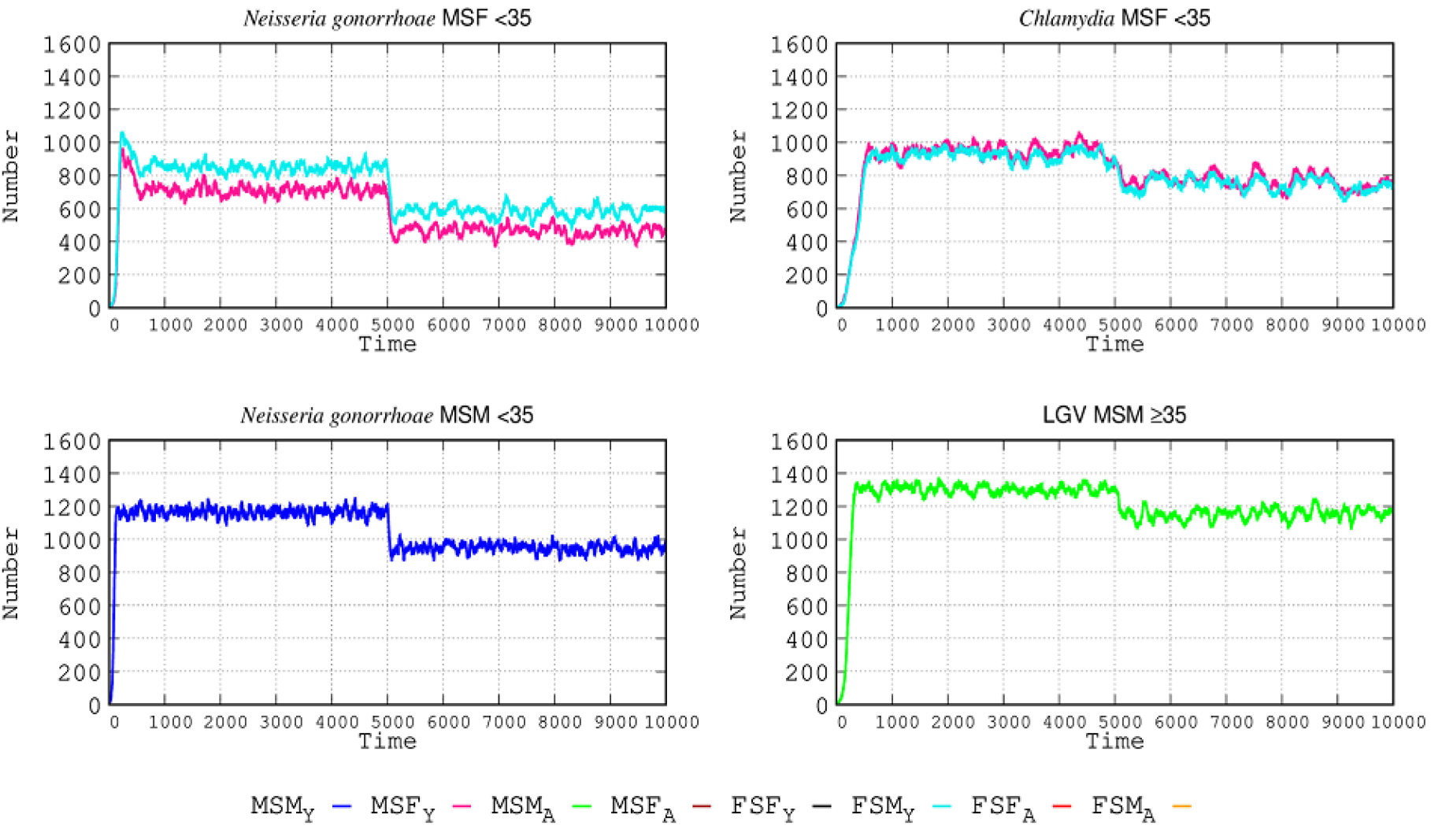
Effect of protective tool use in STI epidemiology. The use of protective tools (such as condoms) was increased (see text for details) in the simulated populations at step 5000, producing a drop in the number of infected cases, of varying intensities for the different STIs. Color lines should be interpreted as in Figure 2.

### The effect of asymptomatic cases in STI epidemiology: the need for a more efficient diagnosis

The most useful epidemiological interventions are based on rapid detection of infection. One of the most significant problems in controlling the transmission of STIs is the high frequency of subclinical (but transmissible) infections, given that the absence of symptomatology impedes interrupting sexual contacts and seeking the etiological agent and specific therapy. Consequently, STIs continue spreading among individuals visiting SEHs. To quantify this effect, we considered the hypothetical case of full diagnosis (no asymptomatic cases), the results of which are shown in Figure 4. For the case of *N. gonorrhoeae*, full awareness of the infection was expected to reduce the number of cases in MSF-FSM <35 by 42% in males and 27% in females. The decrease in number of *N. gonorrhoeae*-infected individuals in the MSM population was much higher at 66%. For *C. trachomatis* infection, the MSF-FSM-infected population <35 could be reduced by 90%, given that this STI is frequently asymptomatic. For LGV in the MSM population ≥35, the number of cases could be reduced by 26%. The reason is that the higher the proportion of asymptomatic cases in a given STI, the more potentially efficacious is the implementation of the diagnosis. With a full diagnosis, the number of *Chlamydia* infections decreases more than the number of LGV cases. This decrease can be explained because *Chlamydia* and *N. gonorrhoeae* infections are both present in the MSF <35 group; when asymptomatic patients are detected, these patients suppress their sexual activity, reducing the number of infections more than in groups exposed to a single STI.

**Figure 4.**
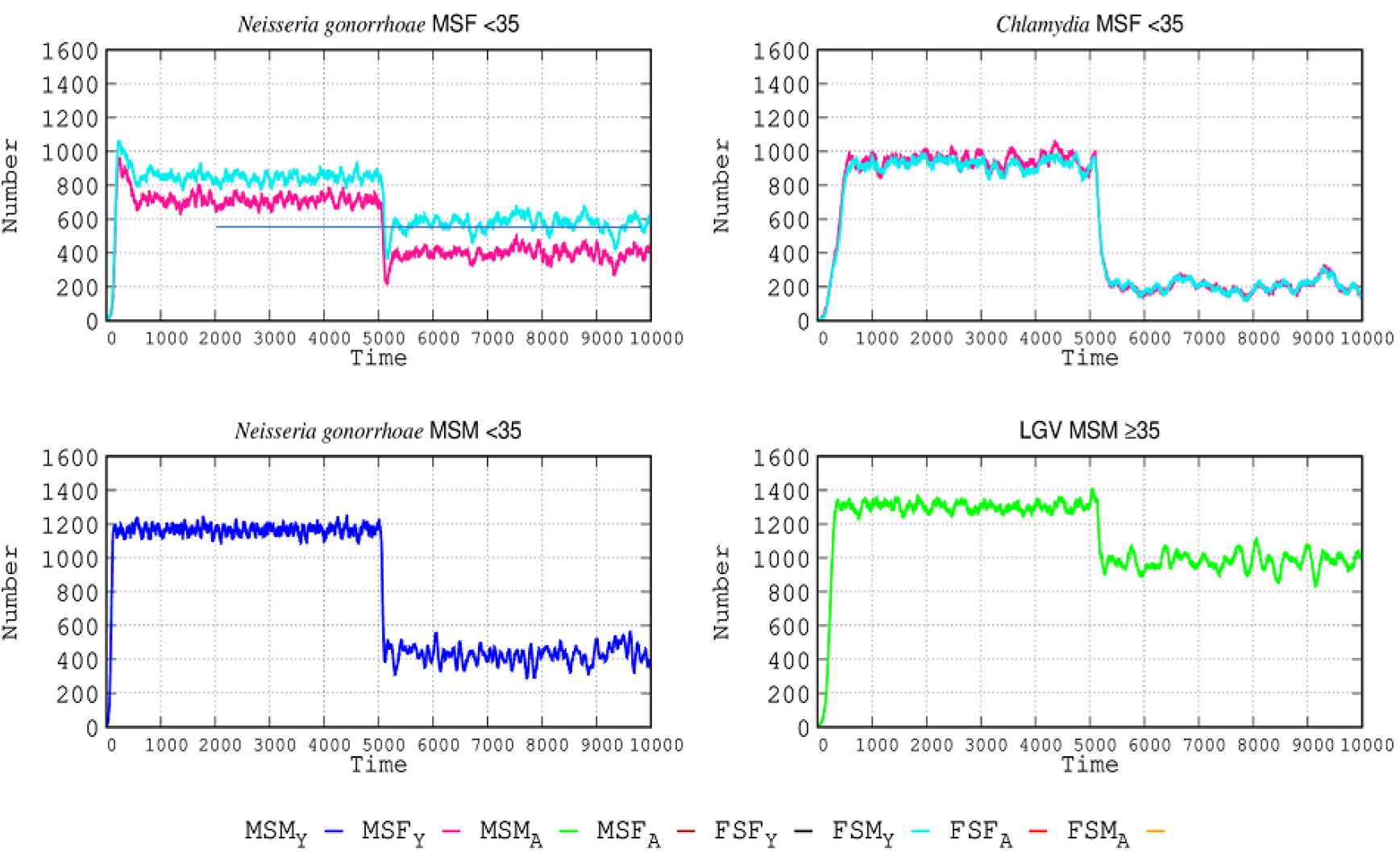
The weight of asymptomatic cases in STI epidemiology. At step 5000, a hypothetical intervention making the detection of asymptomatic cases possible is implemented. In each curve, the left side corresponds to the number of cases with the various STIs in the baseline model study; the right side corresponds to the number of cases if a full diagnosis of infections were possible, detecting and removing asymptomatic cases. Color lines should be interpreted as in Figure 2.

### The effect of sexual contact between populations of different ages in the SEHs

In this variant from the basic scenario, individuals aged <35 and ≥35 years enter into sexual exchanges, with some people <35 having contact with those ≥35 and vice-versa. In this case, each SEH might constitute meeting points for individuals of different ages, contributing to the STI contagion between them. The frequency of trans-age contact is 10% of the movements for those both <35 and ≥35, including equal numbers of MSF, MSM, and FSF. The number of trans-age acting individuals is in fact very scarce in the model, with a total of 8 individuals (4 males and 4 females), and for each gender the 4 types of individuals are represented: MSM or FSF <35; MSM or FSF ≥35; MSF <35; and MSF ≥35. The results are shown in Figure 5. In these trans-age exchanges, individuals from certain groups introduce STIs to other groups, mostly MSF and MSM <35 disseminating *N. gonorrhoeae*; MSF (<35) causing *Chlamydia* contagion; and LGV spreading by those ≥35. We can observe that when 2 different STIs are acquired by individuals of different ages, due to the more promiscuous exchange the absolute number of contagions for a given STI might decrease. The reason is that when in a given age group a new STI is acquired, the number of STIs compete for the same hosts, so that the absolute number of each STI decreases, not the total number of STIs. For instance, when LGV strains enter the MSM <35 population (blue line) after contact with MSM <35 in the ≥35 individuals’ SEH network, the number of *N. gonorrhoeae* infections decreases; similarly, when *N. gonorrhoeae* enters the MSM ≥35 population, LGV contagions decrease (green lines).

**Figure 5.**
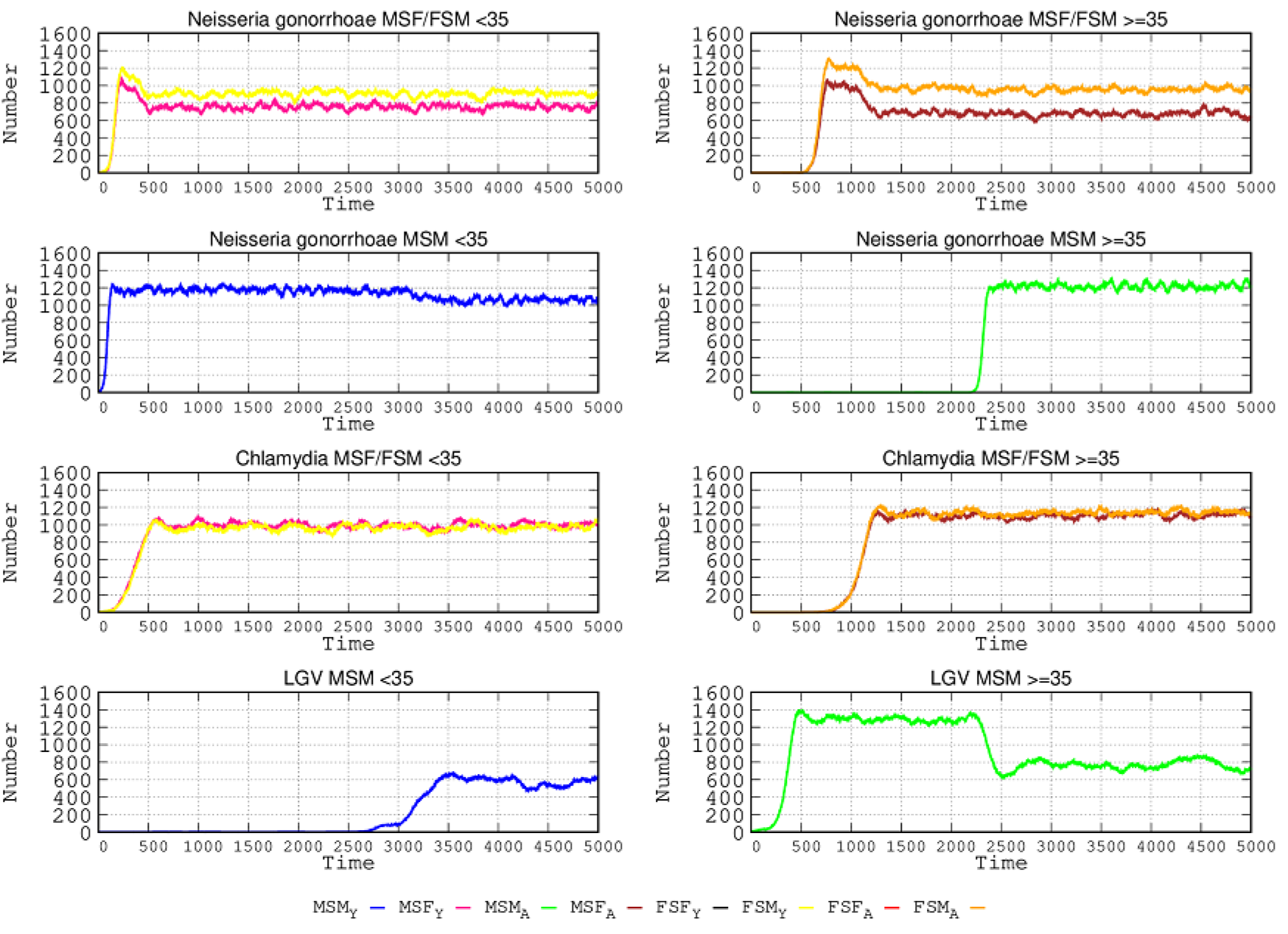
Influence of sexual contact between different age groups in SEHs. Left column: individuals <35; right column, individuals ≥35, visiting SEHs corresponding to ≥35 and <35, respectively. In each row, either the left or the right panel is the group that starts the infection. The groups initiating the infection in the other age group are, in these examples, for *N. gonorrhoeae*, the groups MSF <35 and MSM <35; for *C. trachomatis*, MSF <35; and for LGV, MSM ≥35.

How do protective measures (such as condoms) influence the spread of STIs in this open scenario, in which individuals belonging to different age groups visit the SEHs that correspond to the other ages? As in the specific section (see above) about the effects of protection in the basic scenario, the model considers a protective probability per sexual encounter of 87.55% when using protection. Results are presented in Figure 6.

**Figure 6.**
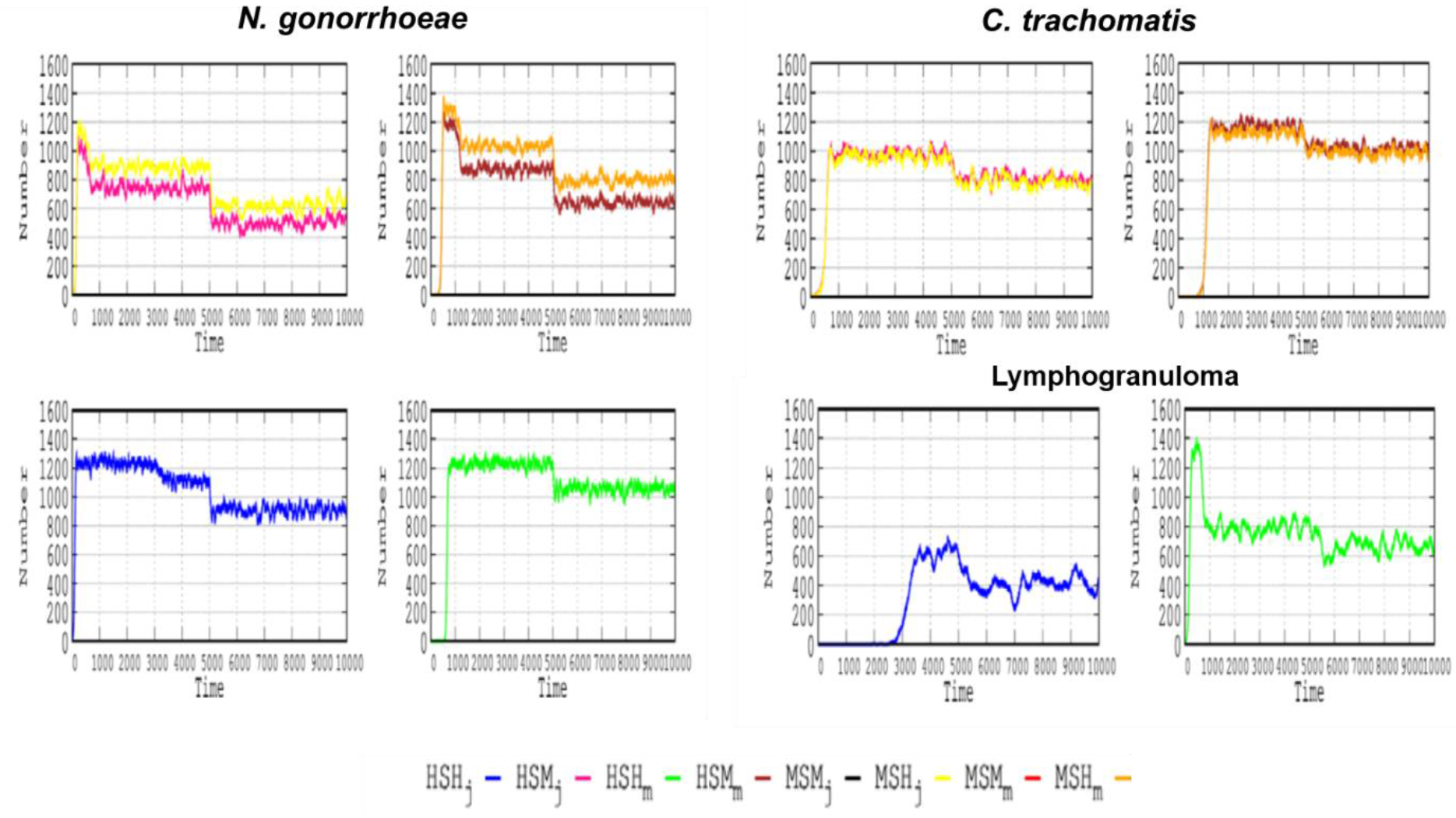
Effect of protection measures in a scenario where individuals in different age groups have sexual relations in SEHs. In each box, the first 5000 steps correspond to the scenario where inter-age interactions are allowed and protection is not applied; during the second steps, protection measures are enhanced. For each STI, the left and right boxes correspond to the populations aged <35 and ≥35.

In this trans-age contact landscape, for the case of *N. gonorrhoeae*, in the <35 MSF population, the decrease of infections associated with protective measures accounted for 44% in males and 33% in females; the protective effect was lower in those ≥35, 40% in males and 26% in females. In the MSM population, protection leads to a lower reduction, a 17% decrease in the number of cases in individuals <35, and of only 14% among those ≥35. For *C. trachomatis* in the MSF population <35, infections were reduced by 19% in both males and females; in the ≥35 group, the reduction was lower, 13% for both genders. In the case of LGV involving MSM, protection provided a reduction of 36% in the <35 group and 13% in the ≥35 group. As noted in a previous section, the benefit of using protective measures is higher in the more transmissible STIs (such as *N. gonorrhoeae*).

The next question to analyze in this scenario of inter-age relations in SEHs is how the complete diagnosis, eliminating non-detected asymptomatic individuals, could affect the number of STI cases. The results of our simulation are presented in Figure 7.

**Figure 7.**
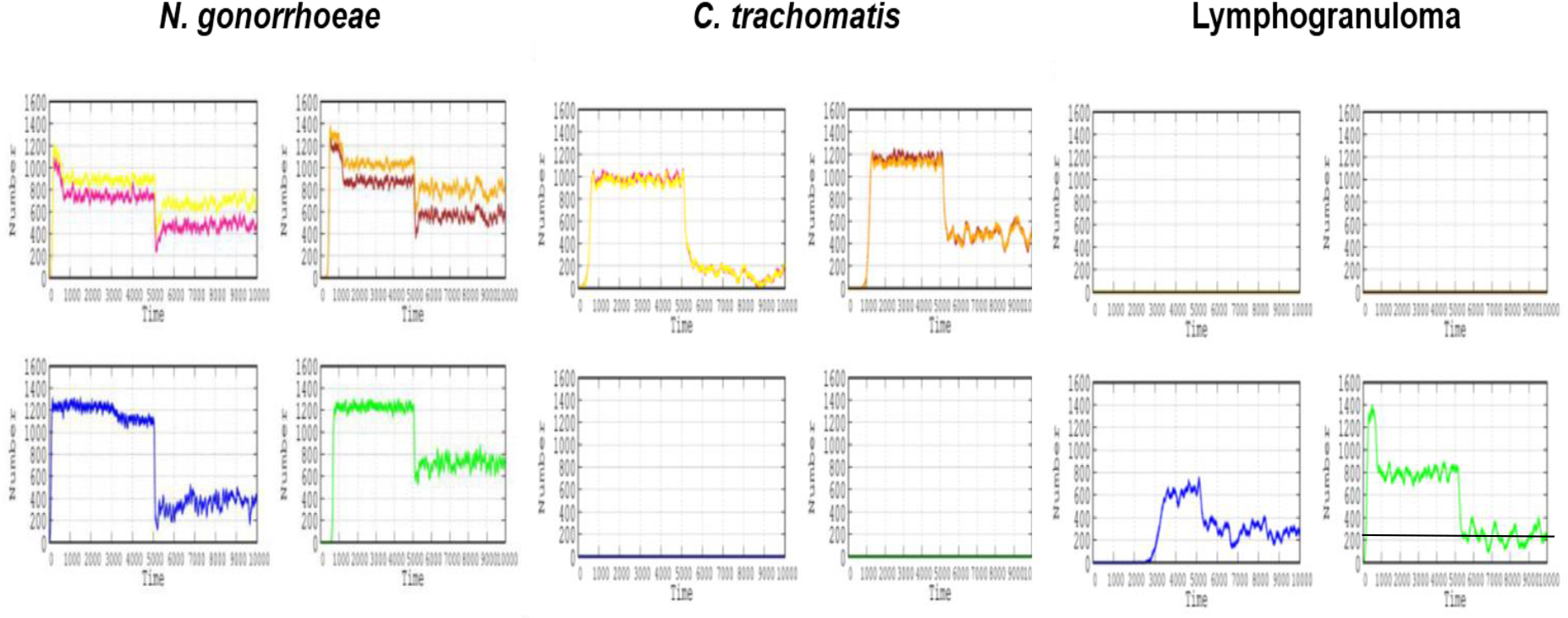
Effect of efficient diagnostic procedures in a scenario in which individuals in different age groups have sexual relations in SEHs. For each STI, the left and right columns correspond respectively to the <35 and ≥35 populations and the MSF (first row) and MSM (second row) populations. Improvement in the efficacy of diagnosing asymptomatic cases is applied at step 5000.

Improved diagnosis of *N. gonorrhoeae* infections has an important but lesser effect than protective tools in MSF populations, potentially reducing the number of cases in 33% and 28% of males and females <35, respectively. In the groups aged ≥35, the reduction is lower: 24% for both males and females. Much more important is the effect of diagnosis in MSM populations, in which the expected reduction is 66% and 52% for the <35 and ≥35 groups, respectively. Our simulation clearly shows that the benefit of detection of asymptomatic patients by applying efficient diagnostic tools could substantially reduce the number of *C. trachomatis* infections, much more than protective measures do. In the MSF population, the number of cases could be reduced 92% for both genders, and to a lesser extent, 58%-56% (males and females) in the <35 and ≥35 age groups, respectively. Lastly, for LGV, precise diagnosis of asymptomatic cases could also be highly beneficial in the MSM population, reducing the number of cases by 52% and 62% in the <35 and ≥35 age groups, respectively. As was shown in the basic scenario, the effect of full diagnosis, eliminating non-detected asymptomatic individuals, is more visible in infections with high rates of asymptomatic cases. Moreover, inter-age movements have increased the number of individuals with more than one STI, and therefore a full diagnosis is more effective.

### The role of the bisexual population

In this simulation, a bisexual male is an MSM-MSF with 50% of his movements visiting MSF or MSM hotspots. For example, visiting hotspots where MSM dominate, and thus able to propagate STIs from MSF or MSM hotspots to other MSF and MSM encounter zones. A bisexual female is an FSF-FSM potentially attending all SEH types, including those where MSF or FSF are more prevalent, and thus able to propagate the STI from MSF and MSM hotspots to MSF and FSF zones. In the simulation presented here, these bisexual persons have an MP-2 movement pattern.

The changes in the present scenario with respect to the basic model (Figure 2) are because STIs cannot be transmitted directly from MSM and FSF populations in the basic model, requiring contagion through the MSF heterosexual population. For instance, in Figure 8, the lymphogranuloma infection originating in MSM enters the MSM-FSM circuit, and from there to the FSF group of individuals. Progressively, STIs start to produce combined contagions in the same type of populations (because of the bridge provided by bisexuals); for instance, *N. gonorrhoeae* and *C. trachomatis* originally infecting the MSF population <35 propagate successively to MSM <35 and later to FSF. In *N. gonorrhoeae*, the introduction of bisexuals in fact increases the number of infections in the MSF group by 8% in females and 10% in males when compared with the basic scenario (Figure 2). Something similar occurs for *C. trachomatis* infection, with increases of 2% in females and 10% in males. No detectable increase in the number of cases occurs among MSM either for *C. trachomatis* or LGV. The most important consequence of the introduction of bisexuals in the SEHs is the contagion of groups that were spared in our basic simulation, such as females in MSF ≥35, and particularly in the <35 FSF circuit, reaching high numbers of infected individuals, particularly by LGV, followed by *C. trachomatis* and *N. gonorrhoeae* (Figure 8).

**Figure 8.**
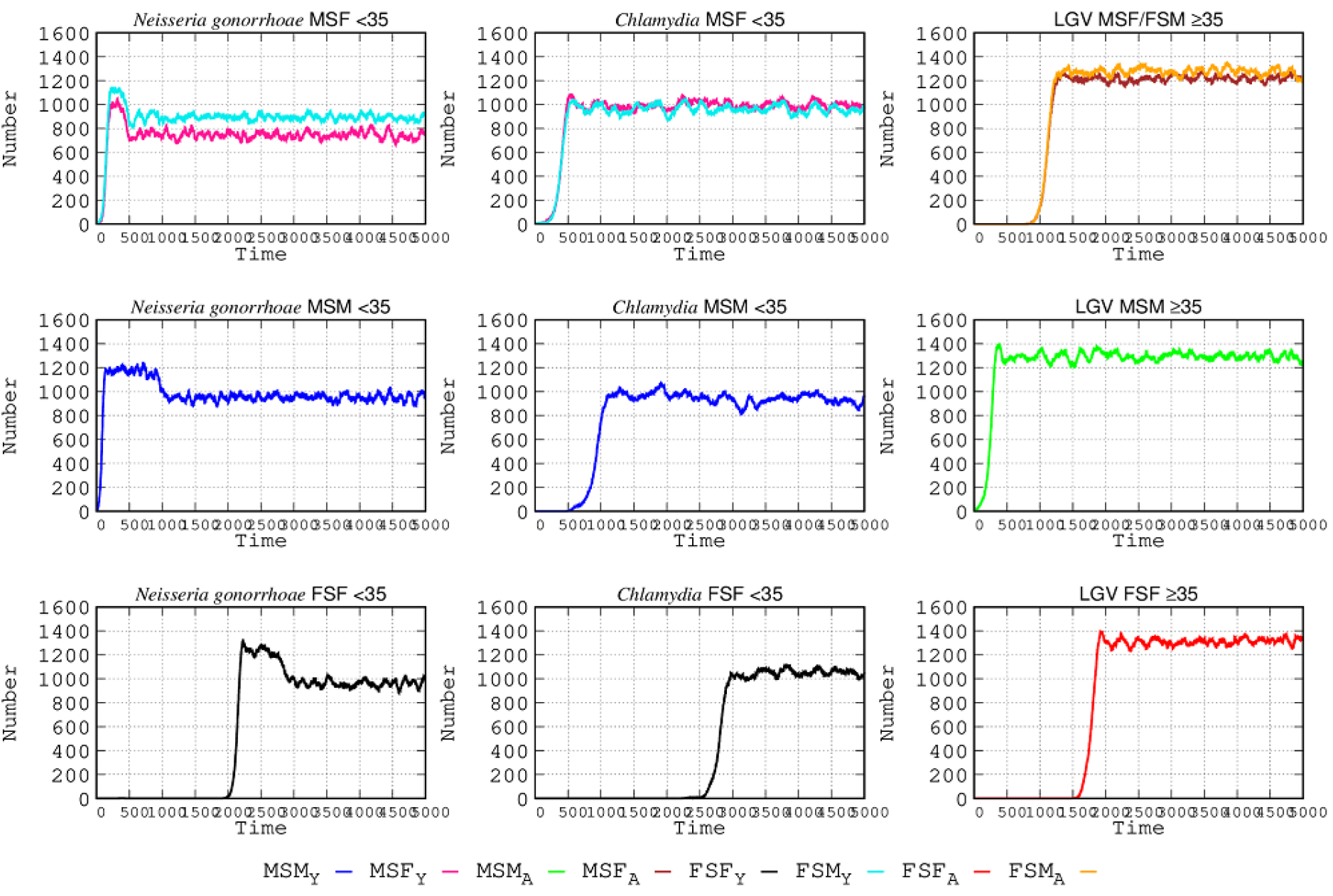
Effect of bisexual shuttlers in STI epidemiology. At the start of the simulation, bisexual individuals primarily belonging to the MSF and MSM <35 population serve to transmit *N. gonorrhoeae*; the MSF population <35 propagates *Chlamydia*; and bisexual MSM ≥35 contribute to LGV contagion.

## DISCUSSION

Social change and its consequent effects on human behavior is a critical factor in the epidemiology of infectious diseases. In our society, the exponential growth of hosts’ mobility is an essential factor contributing to the spread of infectious diseases (20so, 21mk). During the last century, but particularly in recent decades, there might be no better illustration of social change than the sexual revolution (22gj, 23ms). This revolution mostly spread in Western countries, probably grounded in fundamental economic development and consumerism, changes in traditional sexual morality, and its replacement by rights derived from sexual citizenship (the recognition of the individual right to sexual self-determination, also applied to others). All this has been facilitated by the massive and transcultural use of mass media favoring sexual encounters and use of sexual activity enhancers (chemsex), as well as by “security” advances in prevention (PrEP) and STI therapies (24mj, 25bd, 26pr, 27bj, 28fa), as well as the development of hormonal contraception. In our simulation, we have shown how breaking sexual exchange barriers between age groups and regarding sexual behaviors, as in the case of bisexuals, greatly facilitates the spread of STIs in those visiting SEHs.

Progressive societal atomization, based on the reduction of traditional social connections, has been compensated for by the proliferation of multicultural public spaces providing the possibility to enjoy the space with multiple people (29lj). This enhanced mobility strongly facilitates the spread of collective sex and sexual interactions between multiple configurations of gender and age. The cultural expansion of sexual limits is also an expansion in the limits of microbial transmission, and a cause for the dissemination of infectious diseases (30bf).

Our simulation allows us to analyze with an unprecedented level of detail the epidemiological dynamics of STIs when various SEHs are available. It should be noted that the results presented were obtained in a particular “simulation space,” under parameters that can be changed by the program’s user at will. Consequently, the model does not pretend to reflect the general quantitative reality of STIs, but it provides useful information about the relative burden of the various bacterial infections in the different populations of the various sexual orientations visiting SEHs, and regarding the outcomes of adopting interventions. In essence, there are a number of factors that decrease the number of STIs in all studied populations.

First, increasing use of protective measures, such as condoms. This intervention is effective in reducing *N. gonorrhoeae* infection by approximately 35% and 44% in males and females, respectively, in the <35 MSF population. Such reduction is lower (21%) in the MSM population of the same age group. The efficacy in decreasing *C. trachomatis* infections is much lower, decreasing *N. gonorrhoeae* infections in MSF <35 by only 20%. Protection is also limited in reducing the number of cases of LGV in males ≥35, by only 10%. In addition, for both types of *Chlamydia* infections, the application of increased protection takes longer to show an effect than in the case of *N. gonorrhoeae*. Easy availability of protective measures and creating an extended culture of condom use are indeed useful interventions. When MSFs visit a different age-related SEH the protective effect for *N. gonorrhoeae* is increased by 22% and 32% in females and males, respectively, in MSF <35. Similarly, protection lowers the number of cases in the MSM ≥35 population by 11%. In the case of *C. trachomatis* in the MSF population <35, the reduction in number of cases was only 12%, and for those ≥35 only 8%. Lastly, for LGV infection, the number of cases in MSM ≥35 was only reduced by 10% after the increase in protective tools. Low rates of reduction could be associated with more frequent STI circulation with possible reinfections in this particular population. A significant proportion of female LGV infections are oropharyngeal and thus less controllable by condom protection.

Second, improvements in diagnostic screening. The number of STIs is proportional to the number of asymptomatic cases, given that these cases do not reduce or interrupt their sexual contacts since they are unaware of their illness. If we were able to detect all *N. gonorrhoeae* infections early, educating, and treating the patients, the number of cases would be reduced in the <35 MSF population by 42% in males and 27% in females. The decrease in the number of *N. gonorrhoeae*-infected individuals in the MSM population was much higher, at 66%. For *C. trachomatis* infection, cases in MSF <35 could be reduced by 90%, given that this STI is frequently asymptomatic. For LGV in MSM ≥35, the reduction in the number of cases could be decreased by 26%, a proportion compatible with that found by universal testing of rectal *Chlamydia* in MSM (31hy). Recent advances in visualization and rapid procedures for detection (less than 1 hour) of the *ompA* and *orf1* genes of *C. trachomatis* and *N. gonorrhoeae*, respectively, are promising to detect infection without need for sophisticated equipment (32cx). Decentralization of screening procedures resulting from the organization of community points of care could be a useful strategy to implement such diagnostic needs.

Third, the possible future protective role of vaccination and antibiotic prevention therapy (33hb). Although the possibility of delivering a multi-pathogen vaccine against STIs could be a promising approach, it is much more developed for viruses than for bacteria. Recent findings suggest that *N. gonorrhoeae* infection might be prevented with meningococcal B vaccine (34bk). In *C. trachomatis* vaccination, using antigens (most probably MOMP, and/or Pmps, CPAF, and Pgp3), perhaps in combination with adjuvants, might reduce transmission (35dl, 36as). The possible effect of vaccination can be easily tested in our membrane computing model, as we previously did with SARS-CoV-2 (16). Single postexposure dosing of oral antimicrobials, such as doxycycline (and probably macrolides and fluoroquinolones), in high-risk encounters could decrease STI incidence (37mj). However, the benefits versus the risk of antibiotic resistance developing if such a practice is frequently applied should be evaluated. Again, our simulation model could help to assess the appropriateness of such interventions.

Fourth, the role of education to reduce STIs, both for self protection and as a support for the health of the community. Indeed, the all the above-mentioned interventions directed to decrease STIs are fully dependent on greater awareness of the benefits of preventing *N. gonorrhoeae* and *C. trachomatis* infections, which might result, if untreated, in severe complications, including ectopic pregnancy and infertility, fetal infection, and premature delivery (38fc). Certainly, increasing awareness and achieving a change in habits is difficult. Sexual behaviors interact with psychological factors; thus, their modification requires psychosocial interventions (39fu). The possibility that each individual (at least in those visiting SEHs) could have a personal on-line “risk score” (based on behavioral profile, clinical symptoms, and laboratory tests), such as in the form of an interactive computer counseling tool, might be useful (40ms). Such an approach might be combined with “sexual risk behavior scales” (41fe). A condition for the success of such strategies the “risk score” becoming part of social norms (42ka), collectively demanded and accepted by sexual partners and SEH communities at large (43mc). In pursuing such an objective, trends provided by membrane computing simulation, such as those presented in this work, can provide invaluable contributions, including providing material for extracurricular sexual education (44rm).

Lastly, we should be aware that STIs have deeply infiltrated our society, frequently as asymptomatic, hidden, or unsuspected diseases. All health services providing care to the population should recall here the concept of “missed opportunities,” coined by Fauci and Marston when examining the possibilities of controlling sexually transmitted HIV infection (45fa). These opportunities refer to the many encounters between patients with STIs and the health services for any reason (such as in emergency departments) that do not result in a diagnosis (46mz). Knowledge provides opportunities for action. Such knowledge might feed models simulating endo-epidemics in defined landscapes, which could predict the usefulness of targeted interventions.

There are few available mathematical modeling approaches to deal with the complex problem of STI epidemiology (a complex epidemiology involving various pathogens), considering the roles of the various sexual orientations and behaviors among genders and age groups, the hotspots for sexual exchanges and the individual frequentation, the role of asymptomatic patients, the influence of chemsex and PRePs, and the use of protective measures. The standard susceptible-infected-susceptible (SIS) mathematical model was used in the few existing studies (47gg). In fact, the compatibility of competitive exclusion and coexistence of different STIs, also shown in our model, had been proposed long ago using SIS modeling (48cg). Recently, SIS models have incorporated stochastic networks to include asymptomatic individuals and some of the above-mentioned variables, and, as in our model, personal sexual initiatives; the general conclusions are primarily consistent with ours, particularly on the benefits of diagnostic screening and protective measures (49fk). Other modeling studies have used non-linear fractional derivatives with the aim of projecting into the future the possible “maximum peak” of *Chlamydia* epidemics on the basis of cumulative available data in the United States (50vm). However, most of these studies do not reach level of detail of the model presented in this work, showing the possibilities of modeling by membrane computing to allow advances and evaluate interventions in this field.

## Data Availability

All data produced in the present study are available upon reasonable request to the authors.

## ACKNOWLEDGMENTS

In the case of FB, this study was partially funded by the Fundación del Conocimiento Madri+d from the Madrid Autonomous Community through research contracts (AVATAR-EPAMEC) within the Health Start Plus Program of the European Union. For JCG, he was supported by the Instituto de Salud Carlos III (ISCIII), PI20/01397, co-funded by the European Union; and also by the Spanish Network CIBERESP (CB06/02/0053).

## SUPPLEMENTARY MATERIALS

**Table S1.**
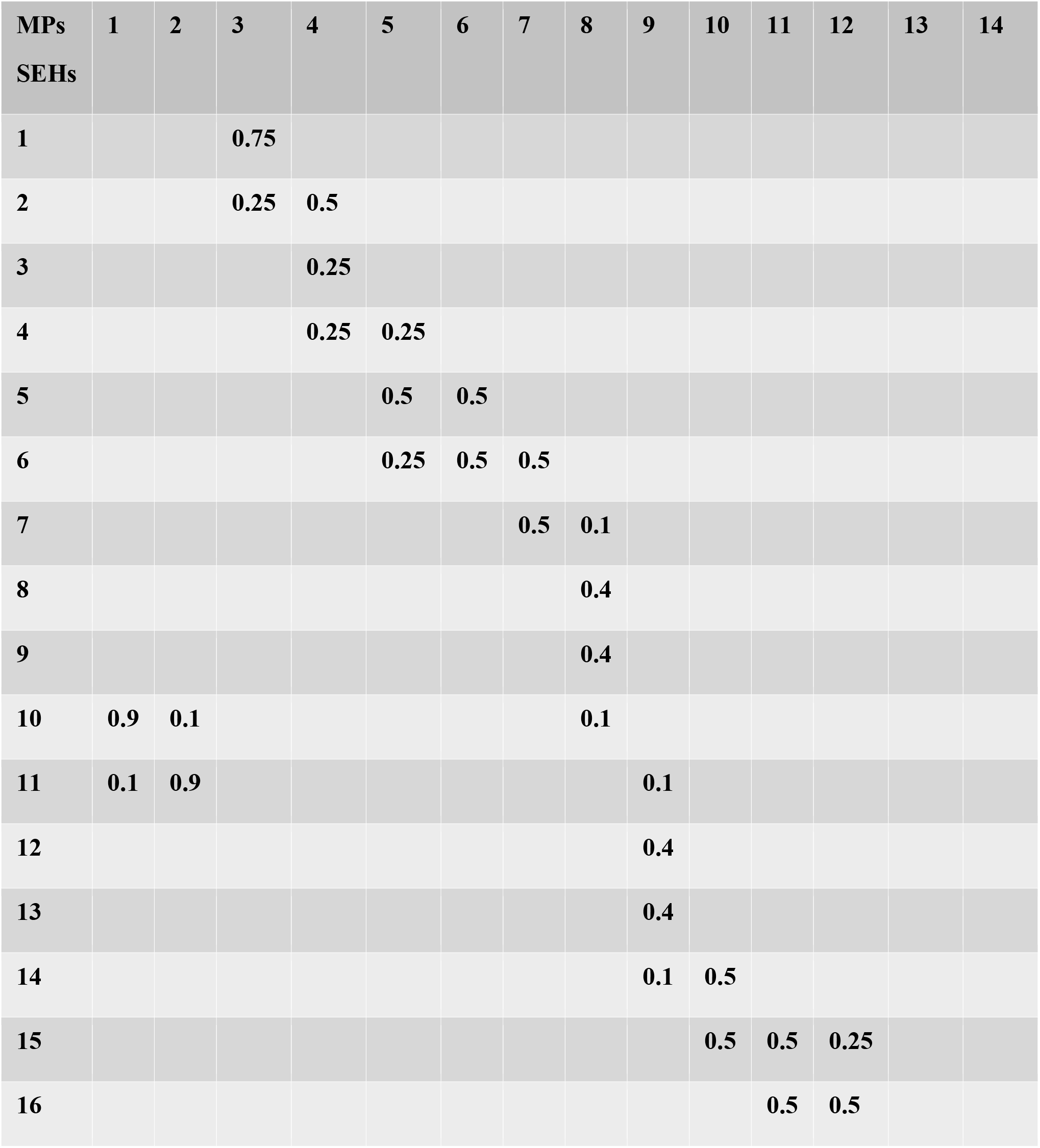
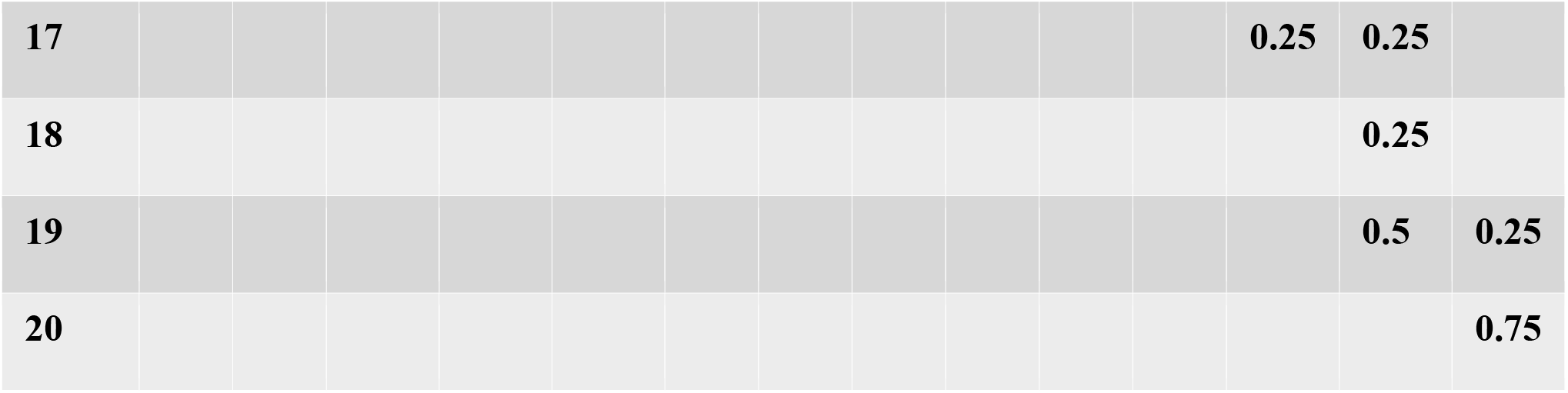
Mobility probability patterns (14 MPs, in columns) to visit sexual exchange hotspots (20 SEHs, in rows) corresponding to the various home residential areas (20 HRAs)

**Table S2.**
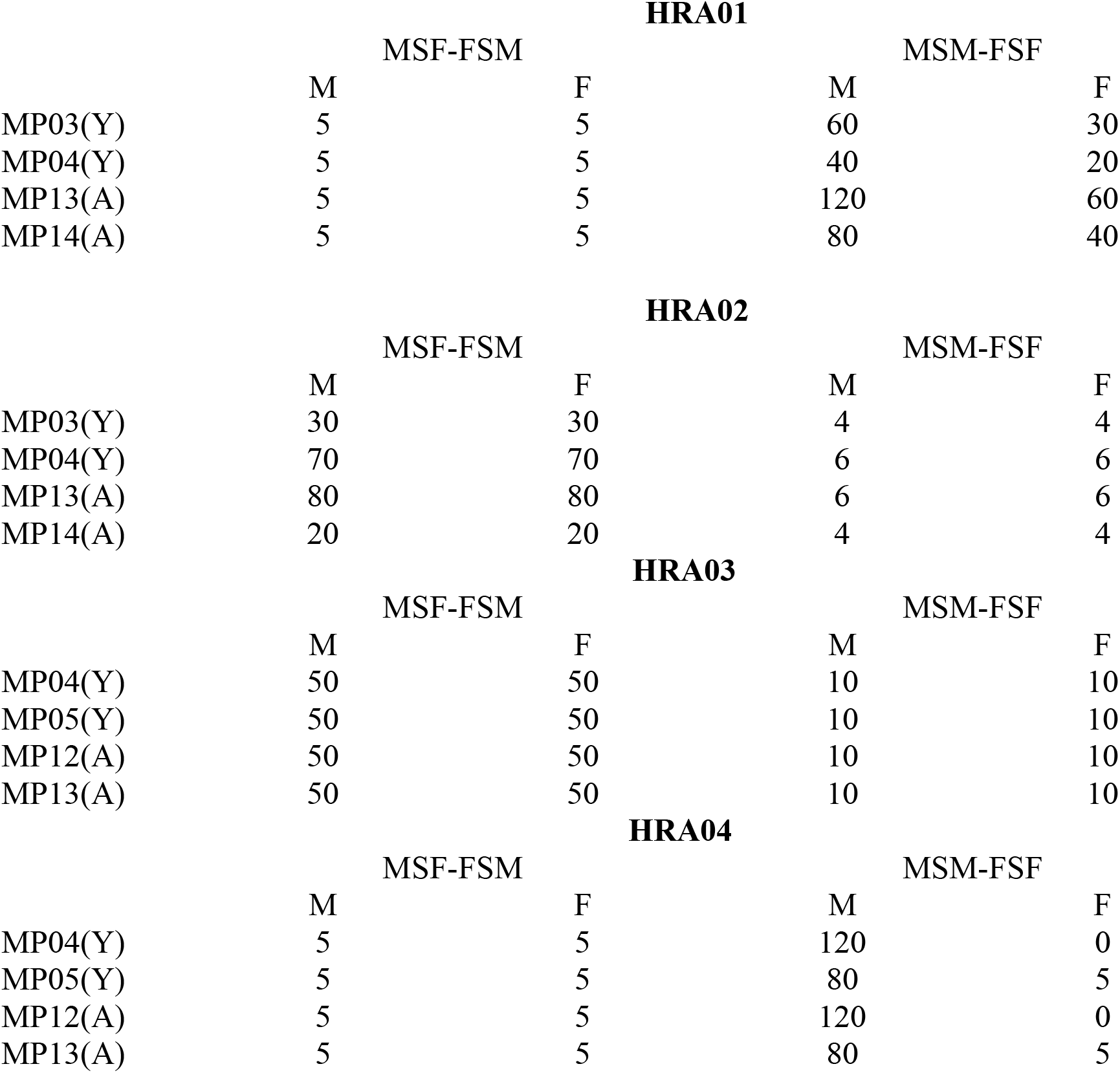
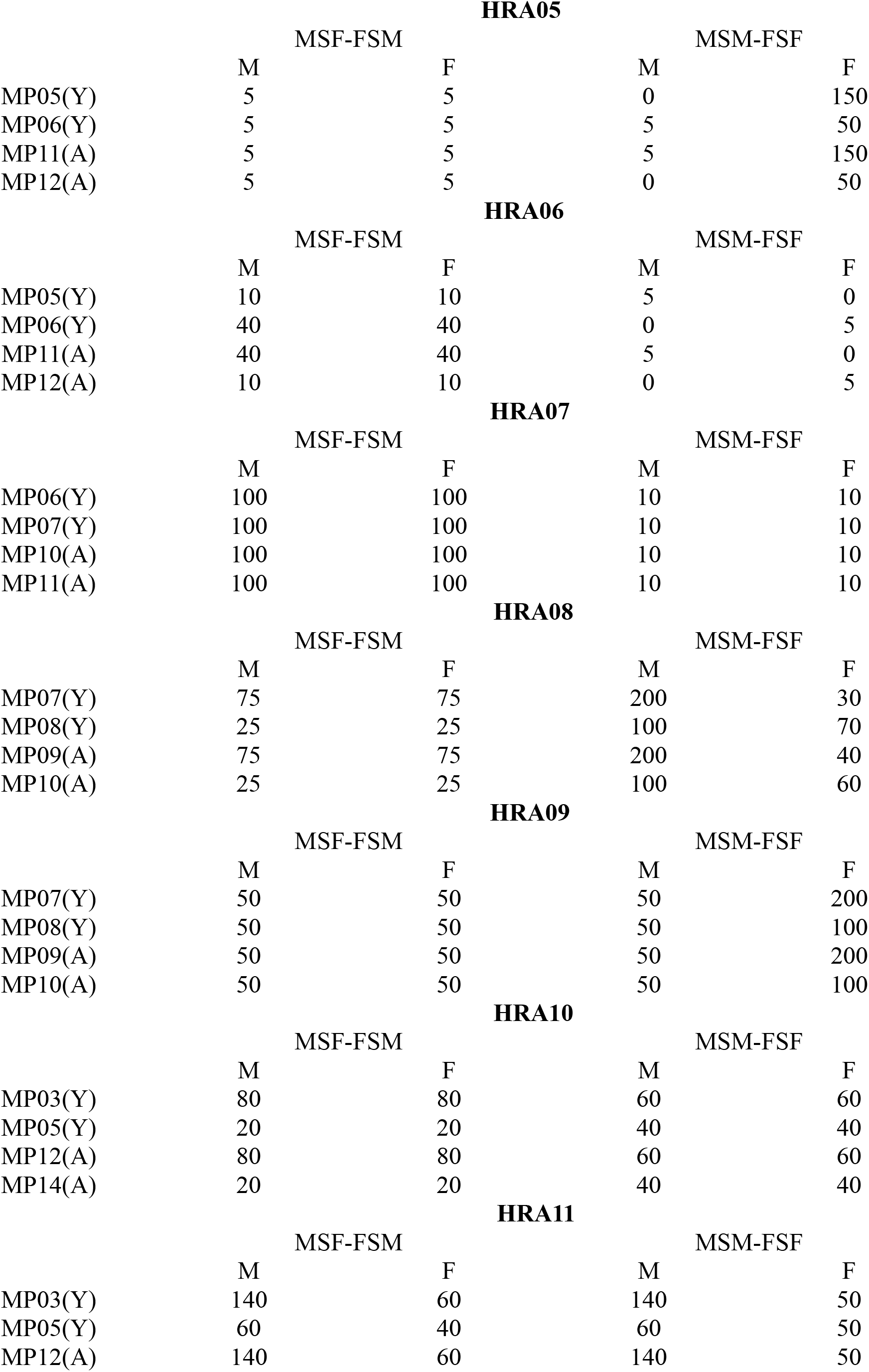
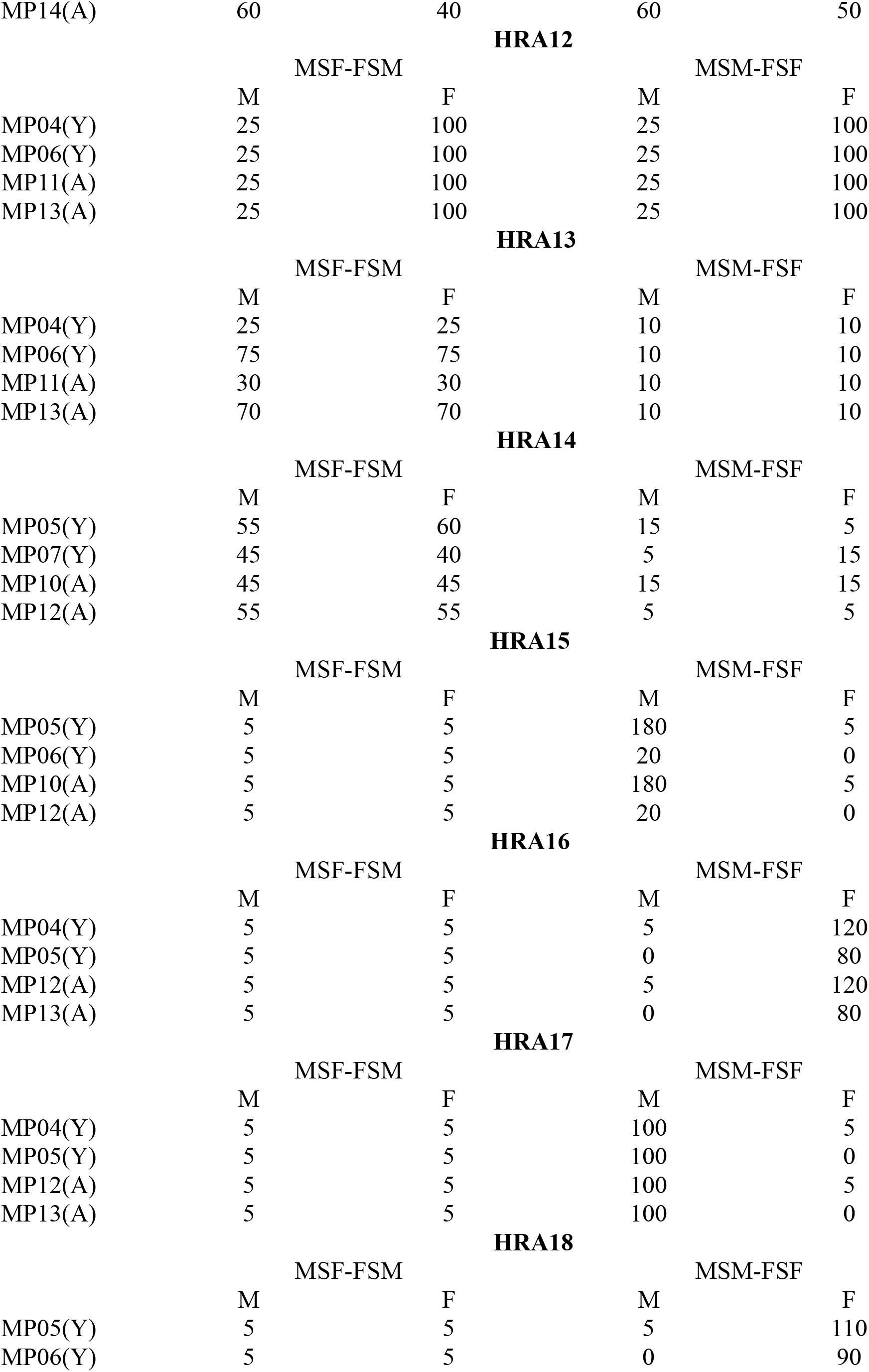
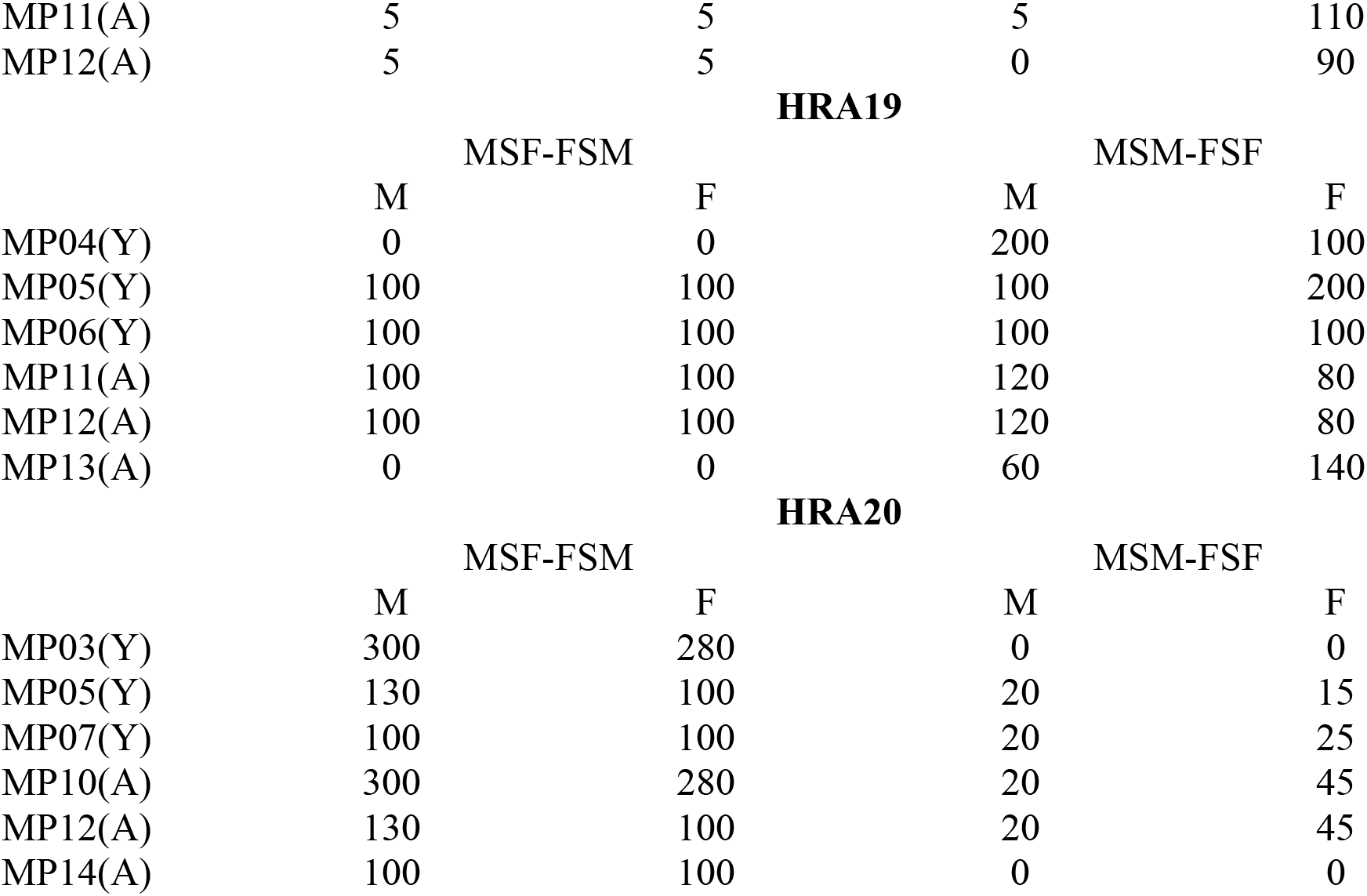
Number of individuals from each of 20 home residential areas (HRAs) visiting 20 sexual hotspots accordingly with 14 mobility patterns (MPs) Example: In the first line of HRA01, MP03(Y) means that in HRA01 there are 5 MSF, 5 FSM, 60 MSM, and 30 FSF, all of them <35 years of age (Y), and all of them following the mobility pattern 3 (MP3)

